# Diagnostic Model of in-Hospital Mortality in Patients with Acute ST-Segment Elevation Myocardial Infarction Used Artificial Intelligence Methods

**DOI:** 10.1101/2020.05.28.20115485

**Authors:** Yong Li

**Author notes:** Corresponding Author:Yong Li, MD, No.2 Clinic, Beijing Anzhen Hospital, Capital Medical University, No. 405,Building No. 5 Madian Nancun, Xicheng District Beijing, 100088 China.

## Abstract

**Background:** Preventing in-hospital mortality in Patients with ST-segment elevation myocardial infarction (STEMI) is a crucial step.

**Objectives:** The objective of our research was to to develop and externally validate the diagnostic model of in-hospital mortality in acute STEMI patients used artificial intelligence methods.

**Methods:** We divide non-randomly the American population with acute STEMI into a training set, a test set,and a validation set. We converted the unbalanced data into balanced data. We used artificial intelligence methods to develop and externally validate the diagnostic model of in-hospital mortality in acute STEMI patients. We used confusion matrix combined with the area under the receiver operating characteristic curve (AUC) to evaluate the pros and cons of the above models.

**Results:** The strongest predictors of in-hospital mortality were age, gender, cardiogenic shock, atrial fibrillation(AF), ventricular fibrillation(VF),third degree atrioventricular block,in-hospital bleeding, underwent percutaneous coronary intervention(PCI) during hospitalization, underwent coronary artery bypass grafting (CABG) during hospitalization, hypertension history, diabetes history, and myocardial infarction history.The F2 score of logistic regression in the training set, the test set, and the validation data set were 0.81, 0.6, and 0.59 respectively.The AUC of logistic regression in the training set, the test set, and the validation data set were 0.77, 0.78, and 0.8 respectively. The diagnostic model built by logistic regression was the best.

**Conclusion:** We had used artificial intelligence methods developed and externally validated the diagnostic model of in-hospital mortality in acute STEMI patients.

We registered this study with WHO International Clinical Trials Registry Platform (ICTRP) (registration number: ChiCTR1900027129; registered date: 1 November 2019).

## Introduction

In the United States, an estimated 605,000 acute myocardial infarction (AMI) events occur each year. ^[1]^In Europe, the in-hospital mortality of patients with ST-segment elevation myocardial infarction (STEMI) is between 4% and 12%. ^[2]^Coronary heart disease, including STEMI, remains the main cause of death. ^[1]^Preventing in-hospital mortality of STEMI is a crucial step. A tool is needed to help early detection of patients with increased in-hospital mortality. The Global Registration Risk Score for Acute Coronary Events (GRACE) can be accessed via mobile devices, so it enjoyed a high reputation among users. Myocardial infarction thrombolysis (TIMI) risk score can predict the clinical manifestations of 30-day mortality in patients with fibrinolytic-eligible STEMI.^[3]^The ACTION (Acute Coronary Treatment and Intervention Outcomes Network) score^[4]^was established in 2011 using 65,668 AMI patients, and 16,336 AMI patients were used to validate as a model for predicting in-hospital mortality. The ACTION model updated in 2016 used more patients and added cardiac arrest as a risk factor.^[5]^Xiang Li used the machine learning method to make a prediction model of in-hospital mortality for STEMI patients. ^[6]^Kwon JM used deep learning to establish a prediction model of in-hospital mortality in STEMI patients, which is better than GRACE score and TIMI score. ^[7]^

The current prediction models had the following problems: People had insufficient understanding of the data set of in-hospital mortality as unbalanced data. The unbalanced data was not converted into balanced data. There was no confusion matrix to be made and the area under the receiver operating characteristic curve (AUC) or C statistic to evaluate the prediction model was not comprehensive. Traditional statistical methods were difficult to deal with the above problems calmly; artificial intelligence methods were needed.

The objective of our research was to develop and externally validate the diagnostic model of in-hospital mortality in acute STEMI patients used artificial intelligence methods.

## Methods

The training dataset was 44,996 patients with acute STEMI from January 2016 to December 2016 in the United States. The test dataset was 43,581 hospitalized patients with acute STEMI from January 2017 to December 2017 in the United States.The validation data set came from 40,498 hospitalized patients with acute STEMI from January 2018 to December 2018 in the United States. Data from the National (Nationwide) Inpatient Sample (NIS) were used for this study.

Inclusion criteria: 1. all those STEMI patients who were hospitalized; 2. all those STEMI patients over 18 years of age.Exclusion criteria: none.

It was a retrospective analysis and informed consent was waived by Ethics Committee of Beijing Anzhen Hospital Capital Medical University. Outcome of interest was in-hospital mortality. All in-hospital mortality was defined as cardiogenic or non-cardiogenic death during hospitalization. The presence or absence of in-hospital mortality was decided blinded to the predictor variables and based on the medical record.

We selected 14 predictor variables according to clinical relevance. Fourteen potential candidate variables were age, gender, cardiogenic shock, atrial fibrillation(AF), ventricular fibrillation(VF),first degree atrioventricular block,second degree atrioventricular block,third degree atrioventricular block,in-hospital bleeding, underwent percutaneous coronary intervention(PCI)during hospitalization, underwent coronary artery bypass grafting (CABG) during hospitalization, hypertension history, diabetes history, and myocardial infarction history. All of them based on the medical record and blinded to the predictor variables. AF was defined as all type of AF during hospitalization. VF was defined as all type of VF during hospitalization. In-hospital bleeding was defined as all type of bleeding during hospitalization.

We kept all continuous data as continuous and retained on the original scale. We used univariable and multivariable logistic regression models to identify the correlates of in-hospital mortality. We entered all variables of Tables 1 into the univariable logistic regression. Based on the variables significantly generated by univariate logistic regression, we constructed a multivariate logistic regression model using the backward variable selection method. We used the Akanke information criterion(AIC)and Bayesian information criterion(BIC)to select predictors; It accounts for model fit while penalizing for the number of parameters being estimated and corresponds to using α =0.157. ^[8]^

**Table 1.**
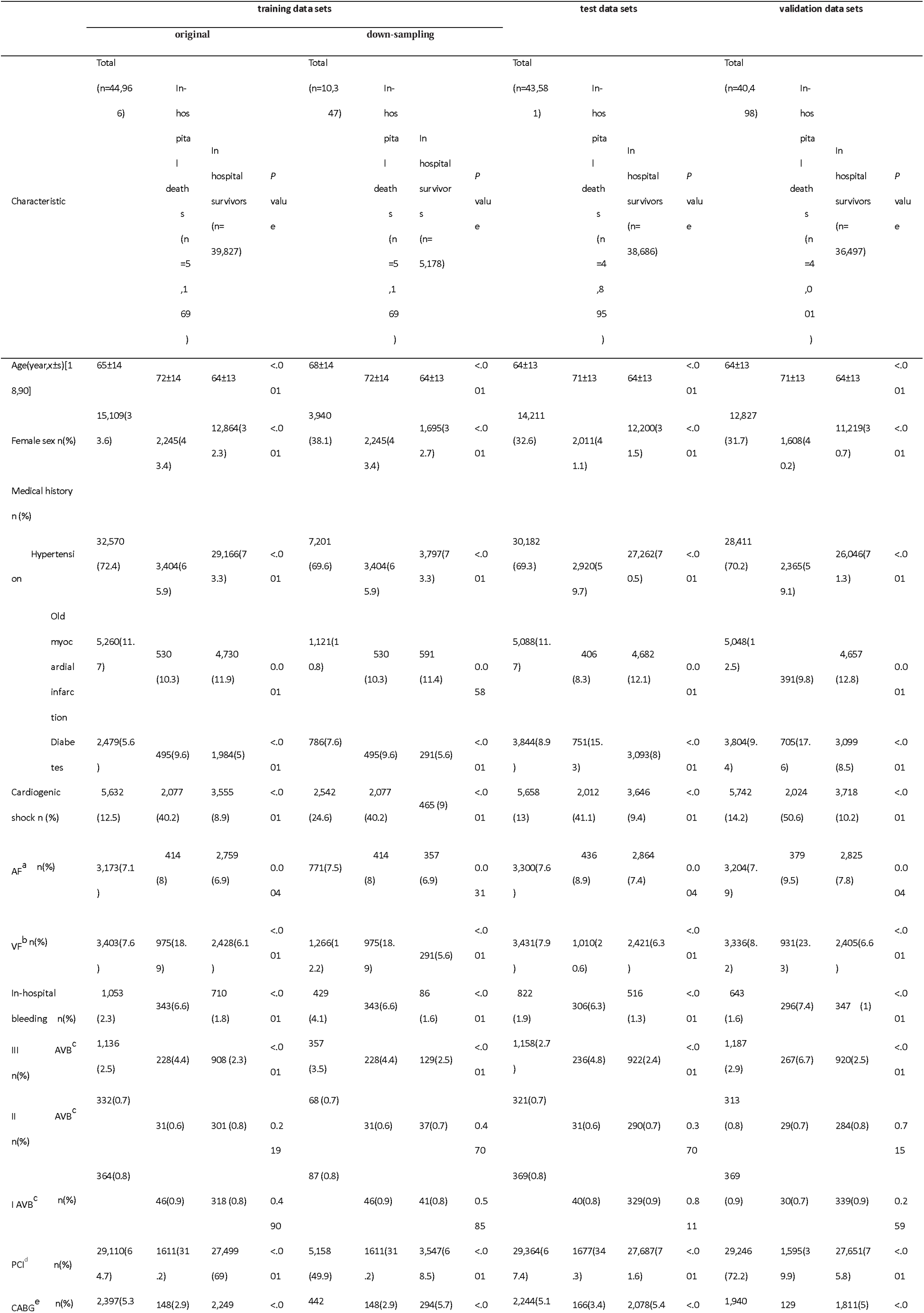
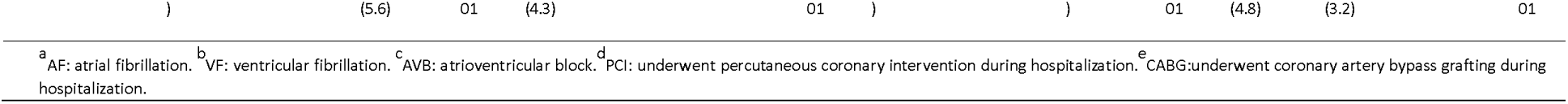
Demographic and clinical characteristics of patient with and without in-hospital mortality

In the training dataset, 5,169 out of 44,966 hospitalized patients (11.5%) experienced in-hospital mortality which represented an imbalanced dataset. We evaluated the effect of common sampling methods including down-sampling methods. Therefore, down-sampling techniques was additionally implemented on the original dataset to create 1 balanced datasets. We randomly selected 13 percent in the survival data as the control group. This ultimately yielded 2 datasets; original, and down-sampling.

To ensure reliability of data, we excluded patient who had missing information on predictors. Discrimination was the ability of the diagnostic model to differentiate between patient with and without in-hospital mortality. This measure was quantified by calculating the AUC^[8]^.

Predictive classifiers were developed based on data from the training set using 5 supervised artificial intelligence methods: (1)Logistic Regression, (2) Random Forest, (3)Extreme Gradient Boosting (XGBoost), (4) K nearest neighbor classification model, and (5)multilayer perceptron. Confusion matrix including Accuracy, Sensitivity, Specificity, Precision, F1 Score, andF2 Score.True Positive=TP,False Negative=FN,False Positive=FP,True Negative=TN.Accuracy = (TP + TN)/(TP + TN + FP + FN), Sensitivity = Recall = TP/(TP + FN), Specificity = TN/(TN + FP), Precision = TP/(TP + FP), F1 Score = 2* Precision * Recall/(Precision + Recall), F2 Score = 5*Precision* Recall/(4*Precision + Recall).F1 Score, was defined as the harmonic average of precision and recall. In addition to F1 scores, F2 scores and F0.5 scores were also widely used in statistics. Among them, in the F2 score, the weight of the recall was higher than the precision, and in the F 0.5 score, the weight of the precision was higher than the recall. The weight of the recall was higher than the precision for the mortality in STEMI patients. We used F2 scorex combined with AUC to evaluate the pros and cons of the above models.

We performed statistical analyses with STATA version 15.1 (StataCorp, College Station, TX). We performed artificial intelligence statistical analysis using Python 3.8.5, Pandas 1.2.1, Sklearn 0.0, Numpy 1.19.2 and Keras 2.4.3.

## Results

The patients’ baseline characteristics of original, and down-sampling were shown in Table 1. Twelve variables (age, gender, cardiogenic shock, AF, VF,third degree atrioventricular block,in-hospital bleeding, underwent PCI during hospitalization, underwent CABG during hospitalization, hypertension history, diabetes history, and myocardial infarction history)were significant differences in the two groups of patient (P <. 157). After application of backward variable selection method, AIC and BIC, all of them remained as significant independent predictors of in-hospital mortality. Results are shown in Table 2. In the test set, 4,895 out of 43,581 hospitalized patients (11.2%) experienced in-hospital mortality. The baseline characteristics of the patients were shown in Table 1. In the validation data set, 4,001 out of 40,498 hospitalized patients (9.9%) experienced in-hospital mortality. The baseline characteristics of the patients were shown in Table 1.By comparing F2 score and AUC of Table 3, we can find that diagnostic model built by dataset of down-sampling was better than those diagnostic model built by dataset of original. By comparing F2 score and AUC of Table 3, we can find that the diagnostic model built by logistic regression was better than those diagnostic model built by decision tree, XGBoost, multi-layer perceptron, and K nearest neighbor. So we used the diagnostic model built by dataset of down-samling and built by logistic regression(“modellog.m).

**Table 2.**
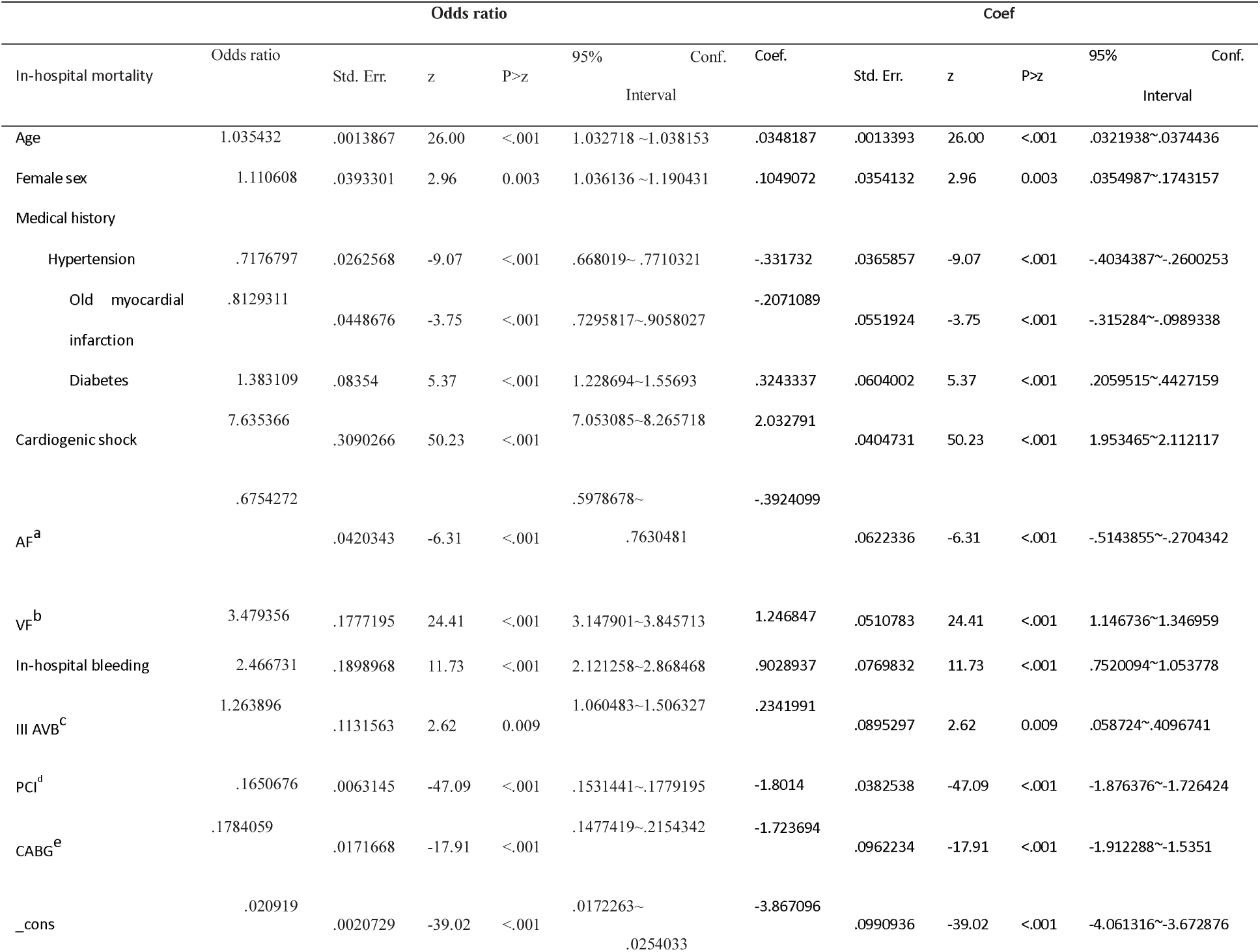
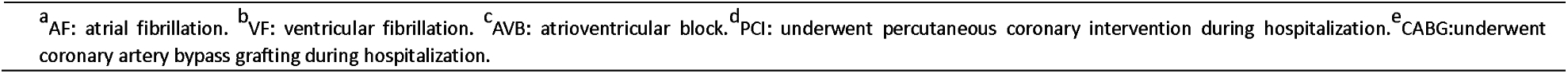
Predictor of in-hospital mortality obtained from multivariable logistic regression modelsin the training data sets

**Table 3.**
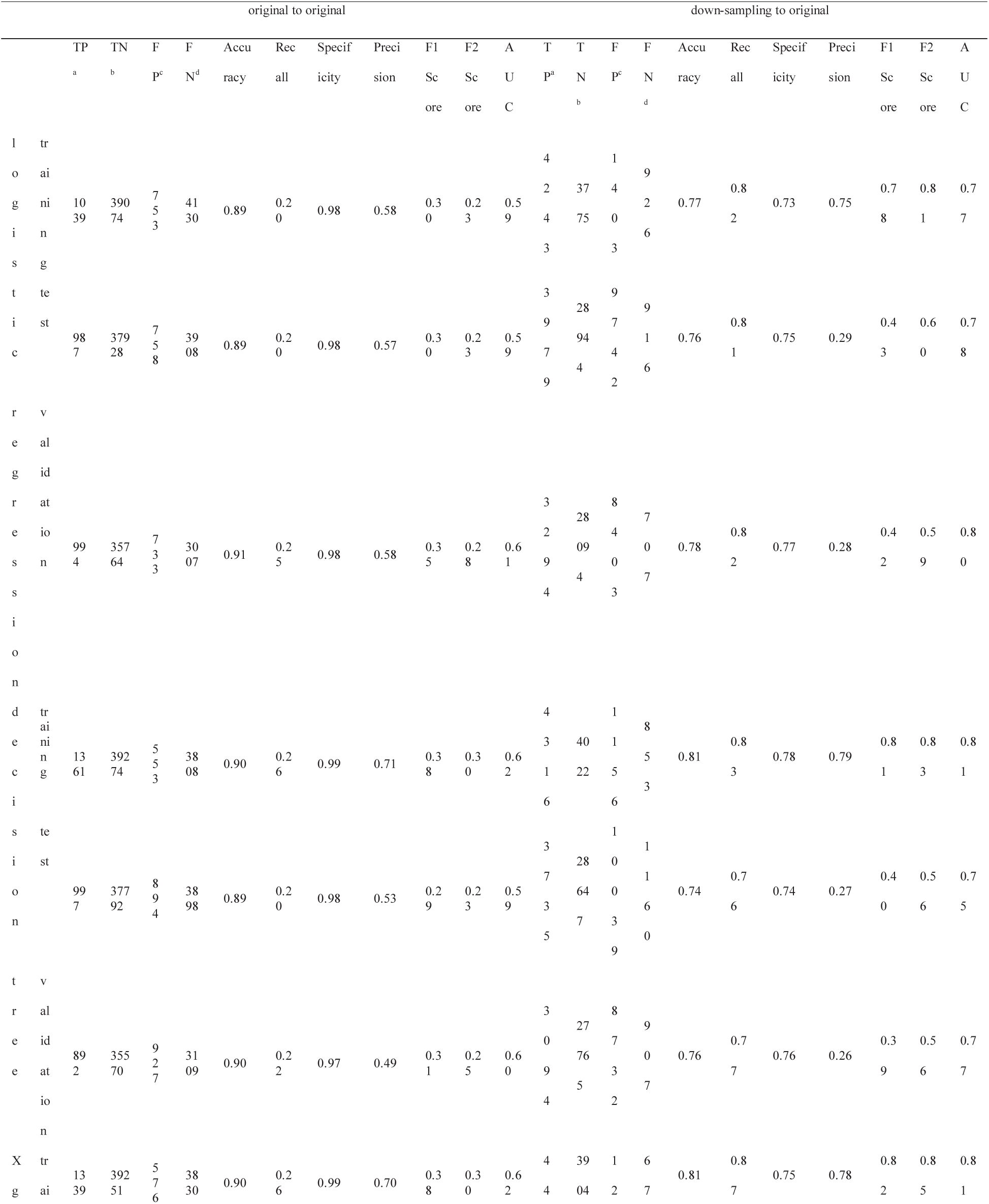
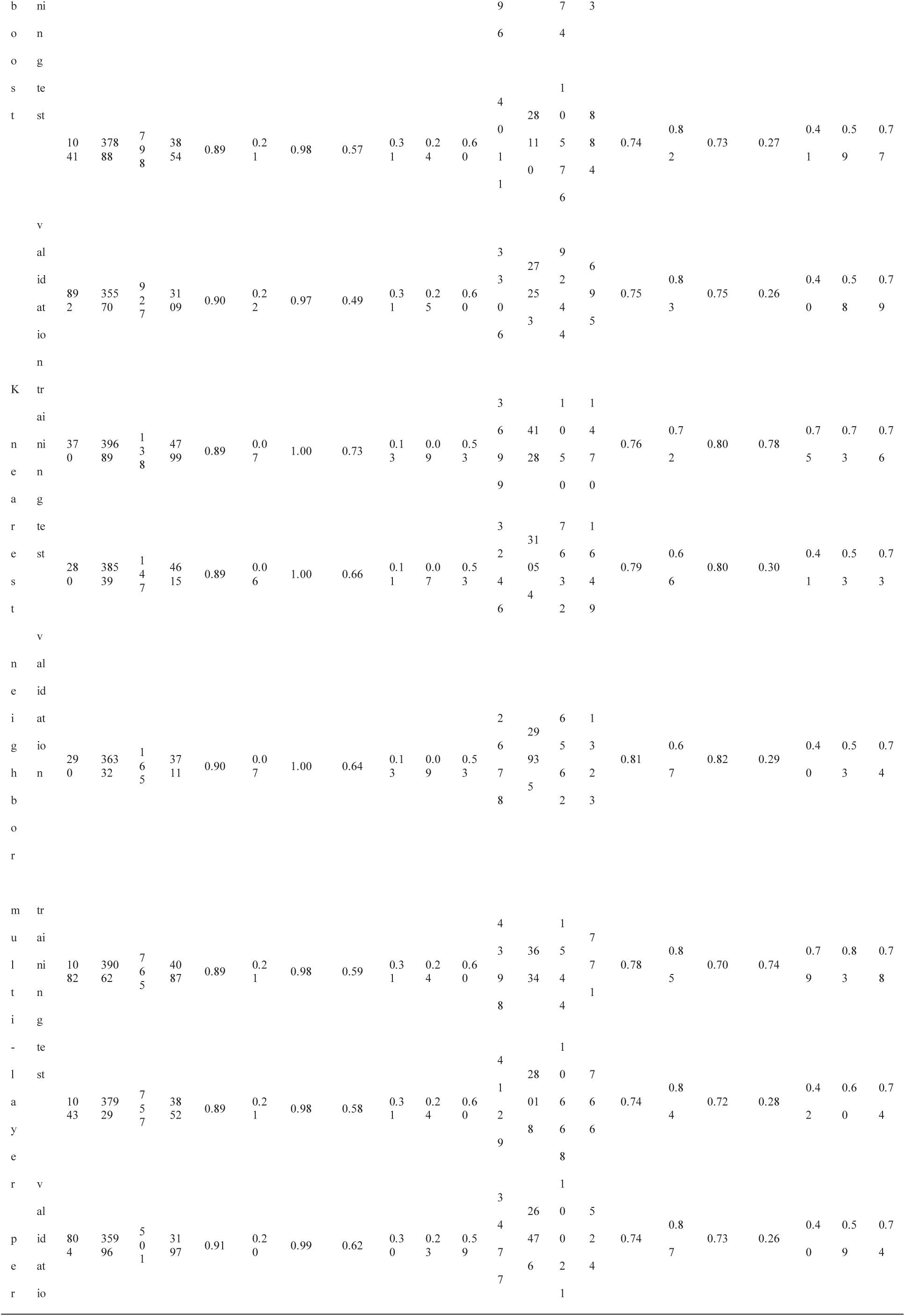
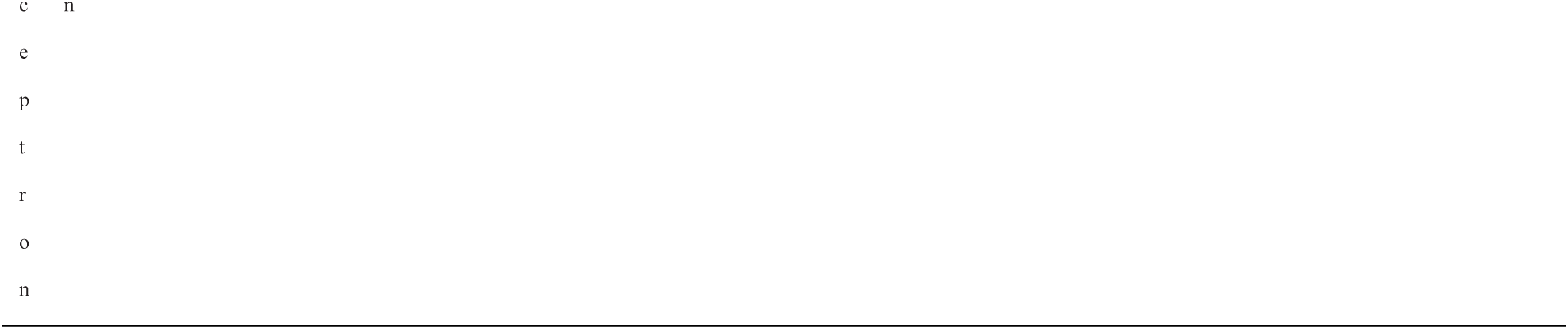
confusion matrix and AUC

The code used for using the diagnostic mode can seen in code1.

If we input the following code on the browser: http://127.0.0.1:8000/ml/predict? AGE=60&FEMALE=1&HBP=1&VF=0&AF=1&OMI=1&CSHOCK=1&IIIAVB=1&DM=1&PCI=1&CABG=0&BLEEDING=1

The result can show below:

[{“features”: {“AF”: 1.0, “AGE”: -1.0, “BLEEDING”: 1.0, “CABG”: 0.0, “CSHOCK”: 1.0, “DM”: 1.0, “FEMALE”: 1.0, “HBP”: 1.0, “IIIAVB”: 1.0, “OMI”: 1.0, “PCI”: 1.0, “VF”: 0.0}, “result”: 0}, {“message”: “1=death,0=alive”}]

## Discussion

In this study, we investigated the predisposing factors of in-hospital mortality in patients with acute STEMI. Age, gender, cardiogenic shock, AF, VF,third degree atrioventricular block,in-hospital bleeding, underwent PCI during hospitalization, underwent CABG during hospitalization, hypertension history, diabetes history, and myocardial infarction history were significant independent predictors of in-hospital mortality.

The F2 score of logistic regression in the training set, the test set and the validation data set were 0.8, 0.6, and 0.6 respectively.The AUC of logistic regression in the training set, the test set and the validation data set were 0.77, 0.78, and 0.8 respectively.The diagnostic model built by logistic regression was the best. So we use the diagnostic model built by logistic regression.

Granger CB et al.observed that age, Killip class, systolic blood pressure, ST-segment deviation, cardiac arrest during presentation, serum creatinine level, positive initial cardiac enzyme findings, and heart rate were independent predictors of in-hospital mortality among 11,389 patients in the GRACE^[9]^. Karen S. Pieper et al. generated the updated GRACE risk model and a nomogram.^[10]^The GRACE risk model has since been upgraded again^[11]^and simplified. ^[12]^TIMI risk score predicting 30-day mortality at presentation of fibrinolytic-eligible patients with STEMI. ^[3]^ C-ACS^[13]^was simple four-variable scores that have been developed to enable risk stratification at first medical contact. ACTION score^[4]^used 65,668 patients to develop and 16,336 patients to validate a model to predict in-hospital mortality. The ACTION model updated in 2016 used more patients (243,440) and added cardiac arrest as a risk factor.^[5]^This was a form of internal validation because their cohorts were randomly created. ^[8]^Xiang Li used the machine learning method to make a prediction model of in-hospital mortality for STEMI patients. ^[6]^Kwon JM used deep learning to establish a prediction model of in-hospital mortality in STEMI patients. ^[7]^

So far, clinicians and researchers usually use GRACE or TIMI scores to guide treatment decisions. Our diagnostic model of in-hospital mortality build upon these studies in several ways. We converted the unbalanced data into balanced data. We used confusion matrix combined with AUC to evaluate the pros and cons of the above models.

Our study has several important limitations including its retrospective nature. The F2 score and AUC of logistic regression in the training set, the test set and the validation data set were modest.

## Conclusion

We had used artificial intelligence methods developed and externally validated the diagnostic model of in-hospital mortality in acute STEMI patients.

## Supporting information

modellog

## Data Availability

The data are demographic, and clinical characteristics of patients with acute STEMI. AGE= age; AF =atrial fibrillation;BLEEDING = in-hospital bleeding; CABG =underwent coronary artery bypass grafting during hospitalization;CSHOCK = Cardiogenic shock; DIED= in-hospital mortality; DM= diabetes history; FEMALE = female;HBP = history of hypertension; OMI=history of myocardial infarction;PCI=percutaneous coronary intervention during hospitalization;VF =ventricular fibrillation. Data from the National (Nationwide) Inpatient Sample (NIS) data from January 2016 to December 2018 in the United States were used for this study. https://www.hcup-us.ahrq.gov/.

## List of abbreviations

AF: atrial fibrillation
AMI: acute myocardial infarction
AUC: area under the receiver operating characteristic curve
FN: False Negative
FP: False Positive
MI: myocardial infarction
NIS: National (Nationwide) Inpatient Sample
ROC: receiver operating characteristic
STEMI: ST elevation myocardial infarction
TN: True Negative
TP: True Positive

## Declarations

## Acknowledgments

Not applicable.

## Statement of Ethics

Ethic committee approved the study. Approved No. of ethic committee: 2019039X.

Name of the ethic committee:Ethics committee of Beijing Anzhen Hospital Capital Medical University. We registered this study with WHO International Clinical Trials Registry Platform (ICTRP) (registration number: ChiCTR1900027129; registered date: 1 November 2019). http://www.chictr.org.cn/edit.aspx?pid=44888&htm=4.It was a retrospective analysis and informed consent was waived by Ethics Committee of Beijing Anzhen Hospital Capital Medical University.

All procedures performed in studies involving human participants were in accordance with the ethical standards of the institutional and/or national research committee and with the 1964 Helsinki declaration and its later amendments or comparable ethical standards. The study was not conducted with animals.

## Consent to publish

Not applicable.

## Availability of data and materials

All data generated or analysed during this study are included in this published article [and its supplementary information files].

## Supplementary materials

The data are demographic, and clinical characteristics of patients with acute STEMI. AGE= age; AF =atrial fibrillation;BLEEDING = in-hospital bleeding; CABG =underwent coronary artery bypass grafting during hospitalization;CSHOCK = Cardiogenic shock; DIED= in-hospital mortality; DM= diabetes history; FEMALE = female;HBP = history of hypertension; OMI=history of myocardial infarction;PCI=percutaneous coronary intervention during hospitalization;VF =ventricular fibrillation. Data from the National (Nationwide) Inpatient Sample (NIS) data from January 2016 to December 2018 in the United States were used for this study.https://www.hcup-us.ahrq.gov/.

## Author Contributions

YL contributed to the generation of the study data, analyzed and interpreted the study data, drafted the manuscript, and revised the manuscript. YL was responsible for the overall content as guarantor. The author read and approved the final manuscript.

