## Supplementary material for "Diagnostic Model of in-Hospital Mortality in Patients with Acute ST-Segment Elevation Myocardial Infarction Used Artificial Intelligence Methods": code1

### import joblib

### import flask

### from flask import Flask, request, url_for, Response

### app = Flask(__name__)

### model2 = joblib.load("modellog.m")

### @app.route("/", methods=["GET"])

### def index():

### with app.test_request_context():

### result = {"predict": {"url": url_for("predict"),

### "params": ["AGE", "FEMALE", "HBP", "VF", "AF", "OMI", "CSHOCK", "IIIAVB","DM","PCI","CABG"," BLEEDING"]}}

### result_body = flask.json.dumps(result)

### return Response(result_body, mimetype="application/json")

### @app.route("/ml/predict", methods=["GET"])

### def predict():

### request_args = request.args

### if not request_args:

### result = {"message": "Please write down the features like as http://127.0.0.1:8000/ml/predict?AGE=80&FEMALE=1&HBP=1&VF=1&AF=1&OMI=1&CSHOCK=0&IIIAVB=0&DM=0&PCI=0&CABG=0&BLEEDING=0"}

### result_body = flask.json.dumps(result, ensure_ascii=False)

### return Response(result_body, mimetype="application/json")

### AGE = float(request_args.get("AGE", "-1"))

### FEMALE = float(request_args.get("FEMALE", "-1"))

### HBP = float(request_args.get("HBP", "-1"))

### VF = float(request_args.get("VF", -1))

### AF = float(request_args.get("AF", -1))

### OMI = float(request_args.get("OMI", -1))

### CSHOCK = float(request_args.get("CSHOCK", -1))

### IIIAVB = float(request_args.get("IIIAVB", -1))

### DM = float(request_args.get("DM", -1))

### PCI = float(request_args.get("PCI", -1))

### CABG = float(request_args.get("CABG", -1))

### BLEEDING = float(request_args.get("BLEEDING", -1))

### vec = [[AGE,FEMALE,HBP, VF,AF,OMI,CSHOCK,IIIAVB,DM,PCI,CABG,BLEEDING]]

### print("vec: {0}".format(vec))

### predict_result = int(model2.predict(vec)[0])

### print("predict_result: {0}".format(predict_result))

### result = {

### "features": {

### "AGE": AGE,

### "FEMALE": FEMALE,

### "HBP": HBP,

# "VF": VF,

# "AF": AF,

### "OMI": OMI,

### "CSHOCK": CSHOCK,

### "IIIAVB": IIIAVB,

# "DM": DM,

### "PCI": PCI,

### "CABG": CABG,

### "BLEEDING": BLEEDING

# },

### "result": predict_result },{"message": "1=death,0=alive"}

### result_body = flask.json.dumps(result, ensure_ascii=False)

### return Response(result_body, mimetype="application/json")

### if __name__ == "__main__":

### app.run(port=8000)
